# The Anomalous Effect of COVID-19 Pandemic Restrictions on the Duration of Untreated Psychosis (DUP)

**DOI:** 10.1101/2024.05.09.24306737

**Authors:** Jessica Nicholls-Mindlin, Hadar Hazan, Bin Zhou, Fangyong Li, Maria Ferrara, Nina Levine, Sarah Riley, Sneha Karmani, Walter S Mathis, Matcheri S Keshavan, Vinod Srihari

**Affiliations:** Medical Sciences Division, University of Oxford, UK; Translational and Clinical Research Institute, Newcastle University, UK; Program for Specialized Treatment Early in Psychosis (STEP), Yale University School of Medicine, New Haven, CT, USA; Yale Centre for Analytical Sciences (YCAS), Yale School of Public Health, CT, New Haven, CT, USA; Institute of Psychiatry, Department of Neuroscience and Rehabilitation, University of Ferrrara, Ferrara, Italy; Beth Israel Deaconess Medical Center and Harvard Medical School, Boston, MA, USA

## Abstract

We investigated the impact of COVID-19 restrictions on the duration of untreated psychosis (DUP). First-episode psychosis admissions (n=101) to STEP Clinic in Connecticut showed DUP reduction (p=.0015) in the pandemic, with the median reducing from 208 days during the pre-pandemic to 56 days in the early pandemic period and subsequently increasing to 154 days (p=.0281). Time from psychosis onset to anti-psychotic prescription decreased significantly in the pandemic (p=.0183), with the median falling from 117 to 35 days. This cohort study demonstrates an association between greater pandemic restrictions and marked DUP reduction and provides insights for future early detection efforts.

## Introduction

In March 2020 COVID-19 was declared a global pandemic^1^. With no vaccine available, restrictions on movement and gathering were used to slow transmission. Despite no evidence for a reduction in the incidence of psychosis, presentations to emergency psychiatric services declined in the first months of the pandemic^1^. In Connecticut, average monthly presentations to one department decreased from 113 to 82 in the pandemic^2^. In a New York City service, there was a 43% decline in emergency psychiatric presentations^3^.

In February 2019, the clinic for Specialized Treatment Early in Psychosis (STEP) in New Haven, Connecticut, completed an early detection campaign ‘Mindmap’^4^, but continued to systematically measure delays in access to treatment following psychosis onset, or the duration of untreated psychosis (DUP). The intrusion of the pandemic presented an opportunity for understanding any association between COVID-19 restrictions and DUP. The clinic measured overall delay between psychosis onset admission, including the time from psychosis onset to first antipsychotic prescription and the subsequent delay from antipsychotic prescription to STEP admission^5^. We hypothesized that COVID-19 related restrictions would be associated with an increase in DUP.

## Methods

### Setting and sampling

The STEP clinic accepts individuals aged 16-35 with non-affective psychosis and DUP <3 years, in 10 towns (population 400,000) around New Haven, Connecticut. All consecutive admissions to STEP between February 1, 2019 and March 21, 2022 were included.

### Exposure

The exposure was COVID-19 related restrictions. The pre-pandemic epoch was defined as the end of the early detection campaign (February 1, 2019) until pandemic onset (March 14, 2020), a time of no restrictions. The early pandemic epoch was the 6-month period that began on March 15, 2020, when stringent restrictions in Connecticut were announced, until September 14, 2020, when most restrictions had been lifted^6^. Subsequent admissions until March 21, 2022, a time of minor restrictions, were included in the late pandemic epoch, which finished 2 years after pandemic onset.

### Measures

DUP-Total was assessed as the time in days between psychosis onset and enrolment into STEP. Components of this overall measure of delay were DUP-Demand (time in days between psychosis onset and first antipsychotic use) and DUP-Supply (time in days between first antipsychotic use and enrolment into STEP). Psychosis onset was dated on the day when the ‘Presence of Psychotic Syndrome’ criteria were met as per the ‘Structure Interview for Psychosis Risk Syndromes’^7^.

### Statistical analysis

Participant characteristics were compared using one-way analysis of variance (ANOVA), Kruskal-Wallis test for continuous variables, and the Chi-square test or Fisher’s exact test for categorical variables. The Wilcoxon rank sum test was used for pairwise comparison between pandemic epochs, due to skew of DUP data distribution. Analyses were carried out with SAS 9.4 (Cary, NC).

### Ethical approval

The authors assert that all procedures contributing to this work comply with the ethical standards of the relevant national and institutional committees on human experimentation and with the Helsinki Declaration of 1975, as revised in 2008. Informed consent was obtained as set out by a protocol approved by Yale University Human Investigation Committee (Protocol Number: 1310012846), which also approves all procedures involving human subjects.

## Results

### Sample characteristics

Participants in the pre-pandemic and pandemic epochs were broadly comparable, except for a higher proportion of American citizens in the late pandemic epoch and more permanent residents in the early pandemic epoch, which is unlikely to affect access to care (Supplementary Table 1). Missing data were minimal, except in household income, which most patients reported ignorance about. Personal income was relatively complete and not significantly different. The sample was young and racially and ethnically diverse, reflecting local demography. The rate of admissions to STEP in the different epochs remained stable (Supplementary Table 2).

### Duration of untreated psychosis

Figures 1a, 1b and 1c show box plots of DUP-Total, Demand and Supply, respectively in the 3 epochs, with p values for pairwise comparisons. Supplementary Table 3 shows descriptive statistics of the three epochs, including ranges. DUP-Total showed a significant reduction from pre-pandemic to early pandemic, with the median falling from 208 to 56 days (p=.0015). This drop appears to be largely accounted for by changes in DUP-Demand, which reduced significantly from a median of 117 to 35 days (p=.183). DUP-Supply did not change significantly but followed the same pattern as DUP-Total with a delay reduction followed by a rebound. DUP-Total significantly rose in the late pandemic compared to the early pandemic (p=.0281), although not reaching pre-pandemic levels.

**FIGURE 1.**
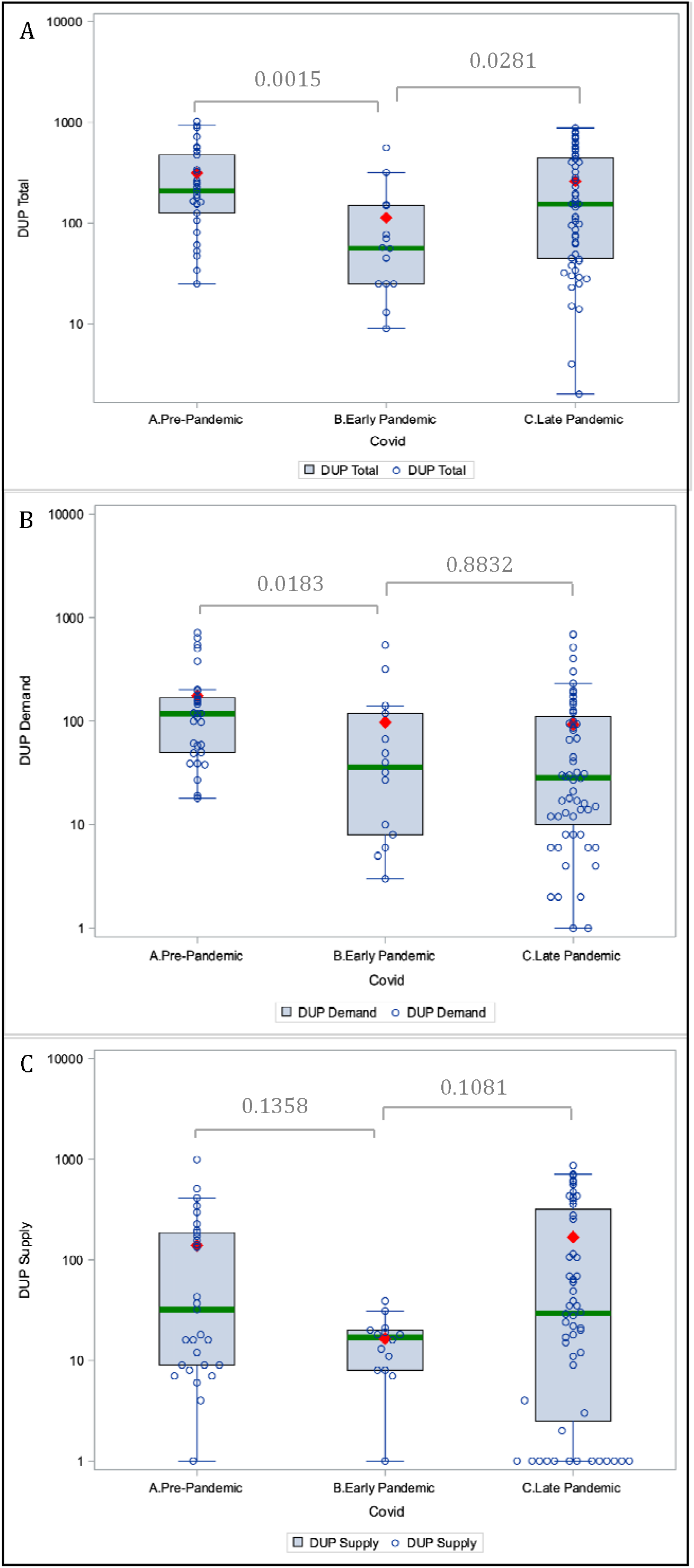
Box and whisker plots of *DUP*-*Total, DUP*-*Demand* and *DUP*-*Supply* in the pre, early and late pandemic epochs. **Legend:** Box and whisker plots of log (DUP) in the pre-, early, and late pandemic epochs for A) *DUP-Total* B) *DUP*-*Demand*, and C) *DUP-Supply*. The green line represents the log (median), the box represents the log (25th and 75th) percentiles, the whiskers represent the maximum and minimum below the upper and lower fence, and the red diamond represents the log (mean). The results of pairwise comparisons of pre-and early pandemic, and of early vs. late pandemic, are reported.

### Summary and conclusions

The overall DUP decreased in the early compared to the pre-pandemic epoch. In the late-pandemic epoch, DUP-Demand persisted at this lower level, while DUP-Total increased. To the best of our knowledge, this is the first report to show a reduction in access delays for psychotic illness during the pandemic. We speculate that this anomalous DUP reduction was caused by earlier awareness by co-habitants of symptoms in the afflicted individual with whom they shared stringent restrictions of movement outside the home. This may have led to quicker help-seeking. Our prior analysis demonstrated that the single largest contributor to DUP in our region was between psychosis onset and first-initiation of help seeking, and that family members were key facilitators^8^. Additionally, reduced patient flow to normally crowded emergency services may have allowed greater attention by clinicians to the identification and management of psychotic symptoms. Both could have led to reductions in DUP-Demand. Since most patients in the catchment receive their first antipsychotic via emergency and inpatient services^8^, this reduction in DUP-Demand is unlikely to be explained by the increasing availability of telehealth services.

The reduction in DUP-Total during the most stringent restrictions and subsequent increase when these were loosened supports a causal role for these restrictions in reducing delays to care. However, DUP-Demand remained low across the two pandemic epochs. This may be related to persistent family availability (e.g. work from home arrangements) to homebound patients, while decays in DUP-Supply may reflect return to pre-pandemic practices.

Some limitations should be noted. The small sample size in the early pandemic group (n=14) increases the likelihood of changes findings. Psychoses are heterogeneous and larger sample sizes are needed to establish if the significant p values are clinically meaningful. Nevertheless, preliminary data from an established early detection service is of interest to other first-episode services, which may observe similar patterns. This data represents the DUP of one region of the country, which may not be generalizable. However, previous analysis of this sample replicated findings of the Scandinavian TIPS study, with an early detection campaign resulting in DUP reduction. Finally, the early detection campaign may have had residual effects on pathways to care, although these would likely have reduced the effects observed in this analysis.

These findings shed light on patterns of presentation to a specialised psychosis service during a global health crisis. Periods of external threat have previously been associated with increased social cohesion^9^; our results could support Durkheim’s theory that major events create group integration^10^, which may promote early detection. It highlights the potentially positive impact of greater attention by household members on reducing delays in care for new-onset psychotic disorders. The magnitude of the reduction in DUP in response to an environmental stimulus is broadly encouraging for early detection efforts that seek to engage observers in facilitating access to care.

## Data Availability

All data produced in the present study are available upon reasonable request to the authors

## Author affiliations

Jessica Nicholls-Mindlin, Medical Sciences Division, University of Oxford, UK. Translational and Clinical Research Institute, Newcastle University, UK.

Hadar Hazan, Program for Specialized Treatment Early in Psychosis (STEP), Yale University School of Medicine, New Haven, CT, USA

Bin Zhou, Yale Centre for Analytical Sciences (YCAS), Yale School of Public Health, CT, New Haven, CT, USA

Fangyong Li, Yale Centre for Analytical Sciences (YCAS), Yale School of Public Health, CT, New Haven, CT, USA

Maria Ferrara, Program for Specialized Treatment Early in Psychosis (STEP), Yale University School of Medicine, New Haven, CT, USA, Institute of Psychiatry, Department of Neuroscience and Rehabilitation, University of Ferrrara, Ferrara, Italy

Nina Levine, Program for Specialized Treatment Early in Psychosis (STEP), Yale University School of Medicine, New Haven, CT, USA

Sarah Riley, Program for Specialized Treatment Early in Psychosis (STEP), Yale University School of Medicine, New Haven, CT, USA

Sneha Karmani, Program for Specialized Treatment Early in Psychosis (STEP), Yale University School of Medicine, New Haven, CT, USA

Walter S Mathis, Program for Specialized Treatment Early in Psychosis (STEP), Yale University School of Medicine, New Haven, CT, USA

Matcheri S Keshavan, Beth Israel Deaconess Medical Center and Harvard Medical School, Boston, MA, USA

Vinod Srihari, (corresponding author) Program for Specialized Treatment Early in Psychosis (STEP), Yale University School of Medicine, New Haven, CT, USA

