## Supplementary Materials for "The Anomalous Effect of COVID-19 Pandemic Restrictions on the Duration of Untreated Psychosis (DUP)"

**Supplementary material**

**Supplementary Table 1**: Demographic characteristics of individuals admitted to the STEP clinic between February 1, 2019, and March 21, 2022, split into COVID-19 epochs ^¥^

|  | **COVID-19 Epoch** | | |  |  |
| --- | --- | --- | --- | --- | --- |
|  | ***Pre-pandemic*** | ***Early pandemic*** | ***Late pandemic*** | **Total** | **P value** |
|  | *N=29* | *N=14* | *N=58* | *N=101* |  |
| **Age at Admission** | | | | | |
| N Missing | 0 | 0 | 0 | 0 |  |
| Mean (SD) | 21 (4) | 23 (4) | 22 (4) | 22 (4) | 0.31 |
| Median (Range) | 20 (16– 35) | 22(18 – 35) | 22 (15 – 32) | 21 (15 – 35) | 0.25 |
| **Years of education completed** | | | | | |
| N (N Missing) | 1 | 1 | 0 | 2 |  |
| Mean (SD) | 12(2) | 13 (1) | 13 (2) | 13 (2) | 0.19 |
| Median (Range) | 12 (9 – 16) | 12(11 – 16) | 13 (9 – 20) | 12 (9 – 20) | 0.23 |
| **First language** | | | | | |
| Missing | 0 | 0 | 2 | 2 |  |
| All Others | 4 (14%) | 1 (7%) | 2 (4%) | 7 (7%) | 0.22 |
| English | 25 (86%) | 13 (93%) | 54 (96%) | 92 (93%) |  |
| **Born in USA** | | | | | |
| Missing | 0 | 0 | 1 | 1 |  |
| No | 5 (17%) | 2 (14%) | 4 (7%) | 11 (11%) | 0.26 |
| Yes | 24 (83%) | 12 (86%) | 53 (93%) | 89 (89%) |  |
| **Household income before taxes, from all sources** | | | | | |
| Missing | 8 | 6 | 5 | 19 |  |
| Less than $40K | 10 (48%) | 3 (38%) | 15 (28%) | 28 (34%) | 0.31 |
| On/above $40K | 10 (48%) | 3 (38%) | 29 (55%) | 42 (51%) |  |
| Unknown/Refused | 1 (5%) | 2 (25%) | 9 (17%) | 12 (15%) |  |
| **Total personal income before taxes, from all sources** | | | | | |
| Missing | 1 | 1 | 2 | 4 |  |
| Less than $40K | 28 (100%) | 13 (100%) | 48 (86%) | 89 (92%) | 0.22 |
| On/above $40K | 0 (0%) | 0 (0%) | 6 (11%) | 6 (6%) |  |
| Unknown/Refused | 0 (0%) | 0 (0%) | 2 (4%) | 2 (2%) |  |
| **Racial background (self-report)** | | | | | |
| Missing | 0 | 0 | 0 | 0 |  |
| White | 8 (28%) | 1 (7%) | 19 (33%) | 28 (28%) | 0.05 |
| Black | 12 (41%) | 11 (79%) | 29 (50%) | 52 (51%) |  |
| Asian | 1 (3%) | 0 (0%) | 1 (2%) | 2 (2%) |  |
| Interracial | 4 (14%) | 0 (0%) | 8 (14%) | 12 (12%) |  |
| Others | 4 (14%) | 2 (14%) | 1 (2%) | 7 (7%) |  |
| **Ethnicity** | | | | | |
| Missing | 0 | 0 | 0 | 0 |  |
| Non-Hispanic White | 6 (21%) | 1 (7%) | 15 (26%) | 22 (22%) | 0.32 |
| Non-Hispanic Black | 12 (41%) | 11 (79%) | 25 (43%) | 48 (48%) |  |
| Hispanic | 4 (14%) | 1 (7%) | 10 (17%) | 15 (15%) |  |
| Non-Hispanic Others | 7 (24%) | 1 (7%) | 8 (14%) | 16 (16%) |  |
| **Citizenship status** | | | | | |
| Missing | 0 | 0 | 1 | 1 |  |
| Permanent Resident | 3 (10%) | 0 (0%) | 0 (0%) | 3 (3%) | **0.01 |
| Refugee | 1 (3%) | 1 (7%) | 0 (0%) | 2 (2%) |  |
| US citizen | 25 (86%) | 13 (93%) | 57 (100%) | 95 (95%) |  |

^¥^rounded to whole numbers*.*

**Supplementary table 2:** Frequency, percentage and rate of individuals recruited to the STEP clinic between February 1, 2014 and March 21, 2022, divided into epochs

| **Epoch (date range)** | **Frequency** | **Percent** | **Cumulative Frequency** | **Cumulative Percent** | **Rate (admissions/month)** |
| --- | --- | --- | --- | --- | --- |
| Pre-campaign  (Feb 1, 2014–Jan 31, 2015) | 23 | 8.46 | 23 | 8.46 | 1.9 |
| Mindmap campaign  (Feb 1, 2015–Jan 31, 2019) | 148 | 54.41 | 171 | 62.87 | 3.1 |
| Post campaign, pre pandemic  (Feb 1, 2019–Mar 14, 2020) | 29 | 10.66 | 200 | 73.53 | 2.1 |
| Early pandemic  (Mar 15, 2020-Sept 14, 2020) | 14 | 5.15 | 214 | 78.67 | 2.3 |
| Late pandemic  (Sept 15, 2020–March 21, 2022) | 58 | 21.3 | 272 | 100 | 3.2 |

**Supplementary table 3:** Mean and median of *DUP*-*Total*, *DUP*-*Demand,* and *DUP*-*Supply* during the pre-, early, and late pandemic epochs^¥^.

|  | **COVID-19 Epoch** | | | **Total** |
| --- | --- | --- | --- | --- |
|  | ***Pre-pandemic*** | ***Early pandemic*** | ***Late pandemic*** |  |
| ***DUP*-*Total* (days)** | | | | |
| N (n missing) | 29 (0) | 14 (0) | 56 (2) | 99 (2) |
| Mean (SD) | 312 (282) | 112 (153) | 260 (262) | 254 (261) |
| Median (range) | 208 (24-1020) | 56 (8-560) | 154 (1-885) | 154 (1-1020) |
| ***DUP*-*Demand* (days)** | | | | |
| N (n missing) | 29 (0) | 14 (0) | 56 (2) | 99 (2) |
| Mean (SD) | 175 (191) | 97 (153) | 92 (154) | 117 (168) |
| Median (range) | 117 (17-714) | 35 (2-541) | 28 (0-690) | 48 (0-714) |
| ***DUP*-*Supply* (days)** | | | | |
| N (n missing) | 29 (0) | 14 (0) | 56 (2) | 99 (2) |
| Mean (SD) | 138 (215) | 15 (10) | 167 (246) | 137 (223) |
| Median (range) | 31 (0-994) | 16 (0-38) | 29 (0-868) | 20 (0-994) |

^¥^rounded to whole numbers.
